# Phase-locked auditory pulse stimulation at home enhancing slow sleep waves: A pilot real-world study

**DOI:** 10.1101/2023.07.02.23292083

**Authors:** Guannan Xi, Xin Zheng, Anchen Gao, Siyang Huang, Huijie Lei, Jing Ding, Qianqian Zhang, Jian Jiang

**Author notes:** Corresponding author: Jian Jiang, Shanghai QuanLan Technology Co., Ltd, No. 3666 Sichen Road, Shanghai.

## Abstract

**Background:** Deep sleep, also known as slow-wave sleep (SWS), is essential for maintaining good health and is characterized by electroencephalographic (EEG) slow-wave activity (SWA). The use of phase-locked auditory stimulation (PLAS) to enhance SWA has emerged as a promising approach. However, the effectiveness of home-based PLAS has not yet been fully established.

**Method:** We used a newly developed wearable EEG device, called LANMAO, to record sleeping EEG signals and synchronize acoustic tones with SWA. We employed a within-subject design to investigate whether acoustic stimulation could increase SWA in a home setting using six subjects (mean age: 29±4.2 years, 3 males). Specifically, we applied acoustic stimulation (STIM) on odd-numbered slow waves and no stimulation (SHAM) on even-numbered slow waves.

**Results:** The PLAS significantly enhanced SWA, theta, alpha, fast spindle and slow spindle activity on STIM condition compare to SHAM condition across all subjects in home setting.

**Conclusion:** These results indicated that the capacity of PLAS, based on LANMAO device, could enhance the SWA in home setting. Our findings shed lights on the wide application of home-based deep sleep intervention.

## Introduction

Slow wave sleep (SWS) is highly associated with crucial functions such as memory consolidation, energy restoration, endocrine regulation, and immune system renew[1, 2]. Low-frequency brain oscillations (0.5-4 Hz), also known as slow-wave activity (SWA), is widely recognized as the hallmark of SWS[3]. Decreasing of SWA has been identified as a transdiagnostic problem across multiple neuropsychiatric disorders such as affective, anxiety, autism and schizophrenia disorders[4–6]. A series of studies have demonstrated that enhancing SWA can enhance memory consolidation, such as declarative and procedural memory[7–9].

Attempts have been made to enhance SWA, including transcranial electrical stimulation[10], transcranial magnetic stimulation[11], and phase-locked auditory stimulation (PLAS)[12]. Among these methods, PLAS has demonstrated the most potential one as a household method, owing to its robust effects and minimal device requirement. The presentation of pulse acoustic stimuli during non-rapid eye movement (NREM) sleep can lead to firing phase synchronization of wide neuronal populations, resulting in SWA increase[13]. However, these studies were normally conducted under strict laboratory conditions, and the reliability in a home setting remains unknown. Although some recent devices claimed to enable closed-loop control in home environment, they either lack supporting evidence or not commercially available yet[14].

In the present study, we utilized a newly developed wearable device to delivery closed-loop stimulation to healthy individuals at their homes. This device consists of two Bluetooth-connected parts: an EEG recorder and a receiver. PLAS was activated by an algorithm deployed in the receiver, which detected SWS stage and the cross-zero time of each slow wave based on the signals from the EEG recorder. By evaluating the effectiveness of PLAS intervention within a home-based context, we provide compelling evidence regarding the feasibility and efficacy of utilizing PLAS in a home setting.

## Method

### Participants

A total of 6 subjects were recruited. The following enrollment criteria were applied: (1) having consistent and regular sleep habits, (2) no history of sleep, psychiatric or neurological disorders, and (3) no physical illnesses that could cause sleep disorders. All participants provided written informed consent. This study was approved by the Ethics Committee of Zhongshan Hospital, Fudan University (Approval No.: SK2020-005).

### Study procedure

Each participant was provided with a LANMAO device (QuanLan Technology Co., LTD, CHINA, as shown in Figure 1A). The device came with pre-set uniform phase-locked algorithm parameters, and after receiving face-to-face instruction, participants were required to wear the device every night before going to bed and choose the paradigm of Closed-loop phase-locked auditory stimulation. When waking up in the next morning, recorder was removed from head. In order to examine the PLAS in real home setting, sleep and awakening times were determined based on individual lifestyle habits, allowing participants to move freely during the night and temporarily leave their bedroom for at most an hour. Additionally, remote assistance was provided by the researchers to help participants address any challenges encountered while using the device.

**Figure 1.**
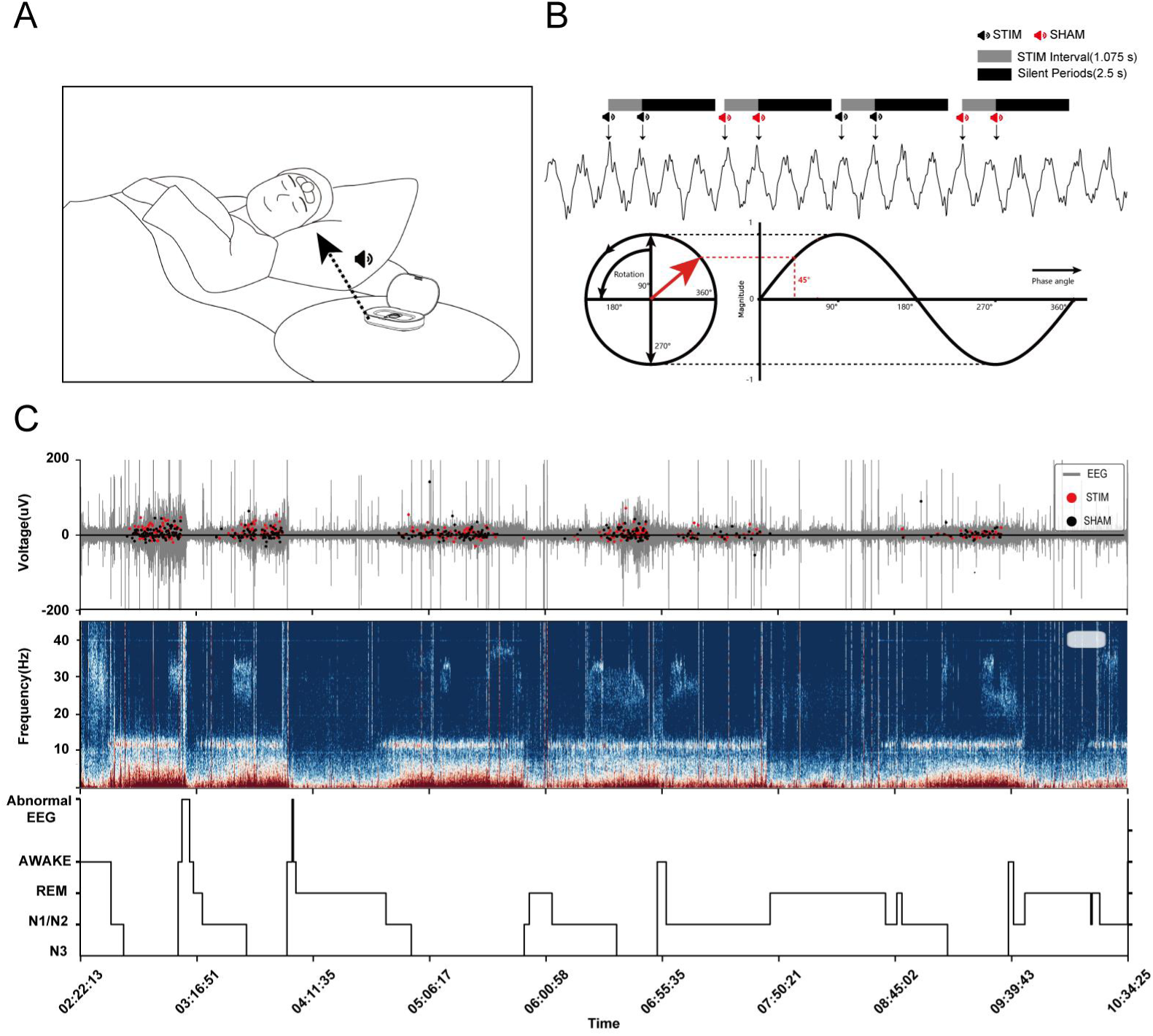
Home setting and the phase-locked stimulation paradigm. (A) The LANMAO device features two components: an EEG recorder (on head) and a receiver (on desktop). The recorder is attached to the participant’s forehead during nighttime sleep, while the receiver received EEG signals, detected slow wave and triggering auditory pulses. The distance between the speaker and the participant’s head is approximately 50 cm, with sound level set at 55 dB. The recorder and receiver were connected via Bluetooth. (B) The stimulation paradigm comprises back-to-back STIM and SHAM blocks, which only get triggered when N3 stages are confirmed by the real-time sleep staging algorithm installed in the receiver. (C) An example of one subject’s overnight home sleep. From top to bottom, EEG waves, spectrogram and hypnogram were showed. In top panel, real stimulations were marked with red dots, and sham stimulation were labeled with black dots.

To minimize the influence of extraneous variables in a realistic home environment and to better evaluate the effectiveness of PLAS intervention, this study employed a within-subject design that did not use separate stimulation and control groups. Instead, we set STIM (stimulation) blocks and SHAM (placebo) blocks for each participant’s night. The STIM blocks (with acoustic stimulation) were assigned to odd-numbered slow waves detected throughout the night, while the SHAM blocks (without acoustic stimulation) were assigned to even-numbered slow waves detected during the entire night (Figure 1B). Each subject was recorded for 1 to 20 days depending on his/her preference. A total of 34 night-sleep EEG data were collected (Table 1).

**Table 1.** Characteristics of participants and number of slow-wave activity (SWA)

| | Mean ± SD |
| --- | --- |
| Age | 29±4.2 |
| Sex (male/total) | 3/6 |
| Days (for each participant) | 20, 3, 2, 1, 1, 7 |
| Number of detected SWAs *(Mean ± SD) | 307±188 |
| *SWAs: slow-wave activities |  |

### EEG recordings device and sleep staging

The LANMAO device recorded whole-night EEG signals near Fp1, with the reference electrode located at the right mastoid (M2). Ag-AgCl hydrogel electrodes were used to keep the impedances below 20 kΩ. EEG Signals were sampled at 500 Hz and then filtered with a band between 0.1 and 100 Hz. Sleep stages were automated classified with the algorithm embedded in the LANMAO device [15] (Figure 1C).

### Detection of slow oscillations and in-phase acoustic stimulation

After sleep stage classification, sleep slow waves during N3 stage were identified based on a previous method. The algorithm was executed on the edge computing platform - LANMAO receiver[15]. Prior to detecting slow oscillation, EEG was pre-processed with a Chebychev second-order bandpass filter featuring cut-off frequencies of 0.5 and 35 Hz.

Once a wave was detected, two audio pulses (100 ms) with a 1.075-second interval were delivered through the speaker in the LANMAO receiver. Then a 2.5-second silent period were performed, in which the slow wave was not detected (Figure 1B). Each acoustic tone was delivered approximately 45 degree ahead of the ascending phase of the wave (Figure 1B), equivalent to about 150 ms during a 0.85-Hz slow oscillation[9]. This accounted for hardware delays and ensured precision audio delivery at the anticipated peak of each SWA up-phase.

The stimulation trials including two types: real stimulation (STIM) and false stimulation (SHAM). In SHAM condition, the slow wave detection was performed as same as the STIM condition, but didn’t play pulse tone.

### EEG preprocessing and analyses

MNE package and custom Python scripts were utilized for pre-processing and analysis of the EEG data.

To evaluate the performance of SO-detection algorithm, we summarized the instantaneous phase of SO when each tone was delivered. Firstly, SO phase was calculated as following processes: EEG signals recorded from the Fp1 region were subjected to band-pass filtering from 0.7 to 1 Hz using a zero-phase shift, Hamming-windowed sinc finite-impulse response (FIR) filter, followed by a Hilbert transform for phase extraction. Secondly, the number frequency of real phase when tone delivering was calculated and plotted as circular histograms for STIM and SHAM conditions, respectively.

Tone-evoked EEG potential was also calculated. Firstly, the EEG signal were band-pass filtered between 0.5 and 35 Hz and averaged across all blocks under STIM and SHAM conditions, respectively. Secondly, EEG data were segmented into 3.5-second clips. Each clip including 0.5-second EEG before stimulation and 3 seconds post-stimulation (STIM or SHAM). Thirdly, each clip was baseline-corrected relative to its 0.5-second pre-stimulus interval. Lastly, the clips were averaged under STIM and SHAM conditions respectively.

Power spectral density (PSD) was computed for each epoch with a frequency resolution of 0.25 Hz (2000-point FFT). The PSD estimates were then averaged across all epochs and smoothed using a three-point moving average [9] under STIM and SHAM condition respectively. To remove the EEG variations across nights and subjects, for each epoch, the power of each 0.25-Hz frequency band was normalized to the cumulative power (MCP) of the 0.25-30 Hz band. Finally, the average percent power change from SHAM blocks to STIM blocks was calculated [9].

The percentage power differences between STIM and SHAM blocks were calculated in following six frequency bands: SWA (0.5-4 Hz), theta (4–8 Hz), alpha (8–12 Hz), slow spindle (12–14 Hz), fast spindle (14–16 Hz), and beta (16–20 Hz). The calculation was performed using the formula: (STIM power - SHAM power) * 100 / SHAM power.

Event-related spectral perturbation (ERSP) plots were used to detect time-varying changes or perturbations in the spectral content that might not been captured in time-averaged signals[16, 17] . The ERSP of each epoch was calculated with and the “mne.time_frequency.tfr_morlet” function from MNE Python package. Briefly, it uses a family of complex Morlet wavelets to decompose signals into time-frequency representations. We used 100 log-spaced frequencies corresponding to each 0.25-Hz frequency bin and increased the number of wavelet cycles used from 1 cycle at 0.8 Hz to 40 cycles at 20 Hz. To estimate phase consistency across trials at different frequencies and characterize the amount of phase-locking across trials, we used inter-trial coherence (ITC). A significant increase in ITC would indicate that EEG activity at a specified time and frequency was phase-locked with respect to stimulus onset.

### Statistical analyses

Differences of tone-delivering phase between STIM and SHAM conditions were examined with Watson-William test. Paired Student’s *t*-tests (two-tailed, even STIM V.S. odd SHAM) were used to determine differences in EEG signals between the STIM and SHAM conditions for all other analyses. The p-values for all tests were adjusted with FDR method. Averaged values were mean ± SEM.

## Results

### Phase-locked tone pulses locked to the upstate of slow oscillation

Phase-locked tone stimulation was applied when slow wave was detected in home setting (figure 1A, B). For each subject, whole-night EEG was recorded and then the sleep stages were categorized based on EEG spectral characters (figure 1C).

To examine the accuracy of phase-locked tone delivery, the tone-locked instantaneous phase of slow oscillation was calculated offline. The tone-locked phase was 30.5 ± 84.6° in STIM condition, while 33.8 ± 81.4° in SHAM condition (Figure 2A). There was no significant difference between the locked phases in two conditions (Watson-Williams circular *t*-test, p=1.0). Additionally, the mean direction of these plots was not significantly different from the target phase of 45 degrees (circular v-test, p=1.0), indicating that the algorithm successfully captured the target phase.

**Figure 2.**
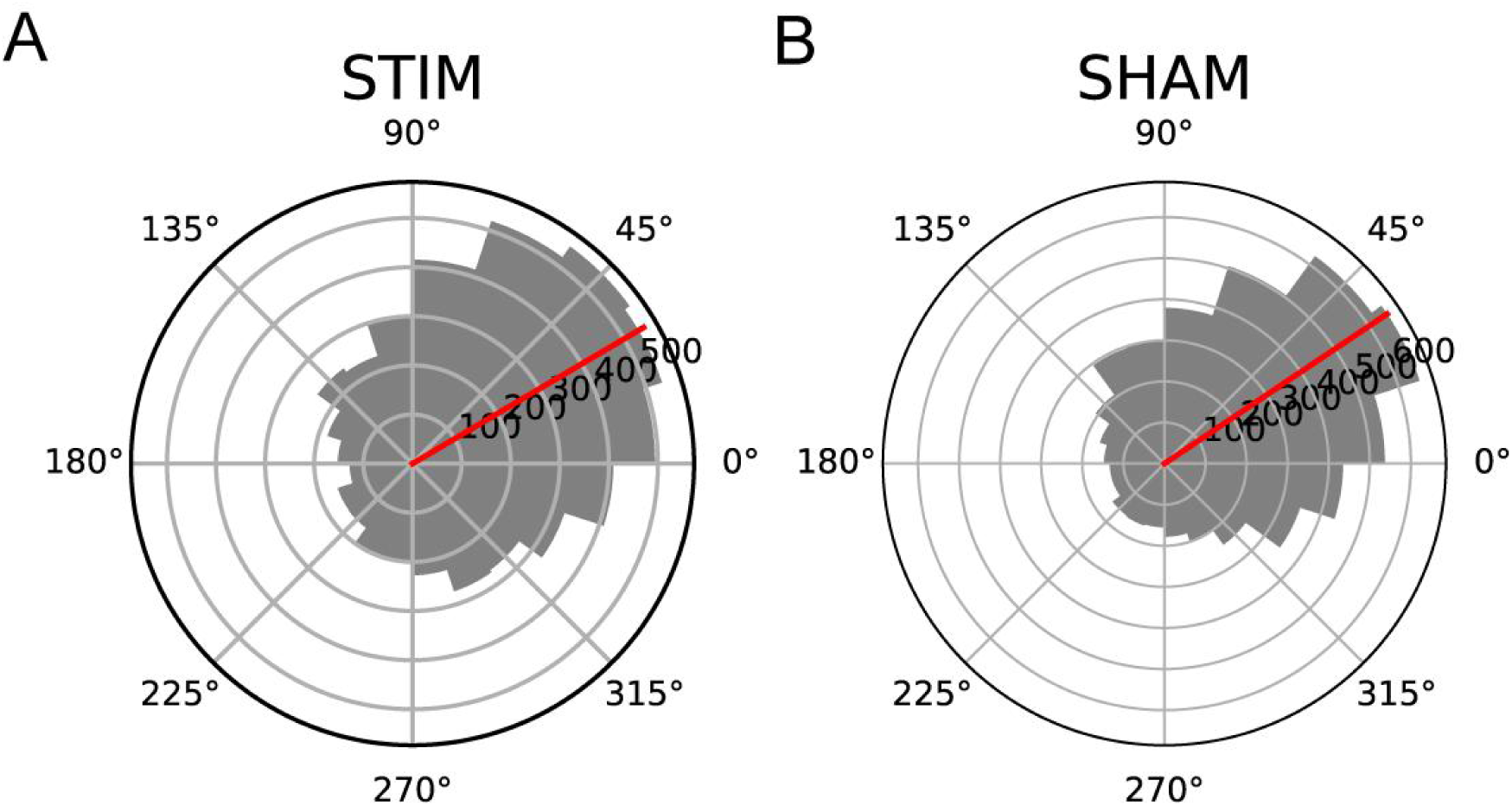
Offline examination of the slow wave phases when audio-playing commands were triggered. The circular histograms(A and B) depict the distribution of the number of tones delivered during the STIM and SHAM conditions for all participants, without considering individual differences. The data collapsed across all participants.

### Phase-locked tone stimulation significantly enhanced low-frequency EEG power

Then we examined whether the sleep EEG activity can be changed by the phase-locked tone stimulations. Power-frequency spectra were calculated and compared between STIM and SHAM conditions (Figure 3). We found that the power of all frequency bands under 20 Hz significantly enhanced in STIM condition (Figure 3A, p < 0.05). The spectra power differences of traditional EEG bands between STIM and SHAM conditions were also calculated. Significant differences were found in SWA (169.5 ± 4.8%), Theta (80.5 ± 1.9%), Alpha (78.0 ± 2.3 %), SS (95.0 ± 3.1%), FS: (69.0 ± 2.5%) and Beta (42.0 ± 2.2%) bands, while not Gamma band (181.5 ± 39.4%) (Figure 3B).

**Figure 3.**
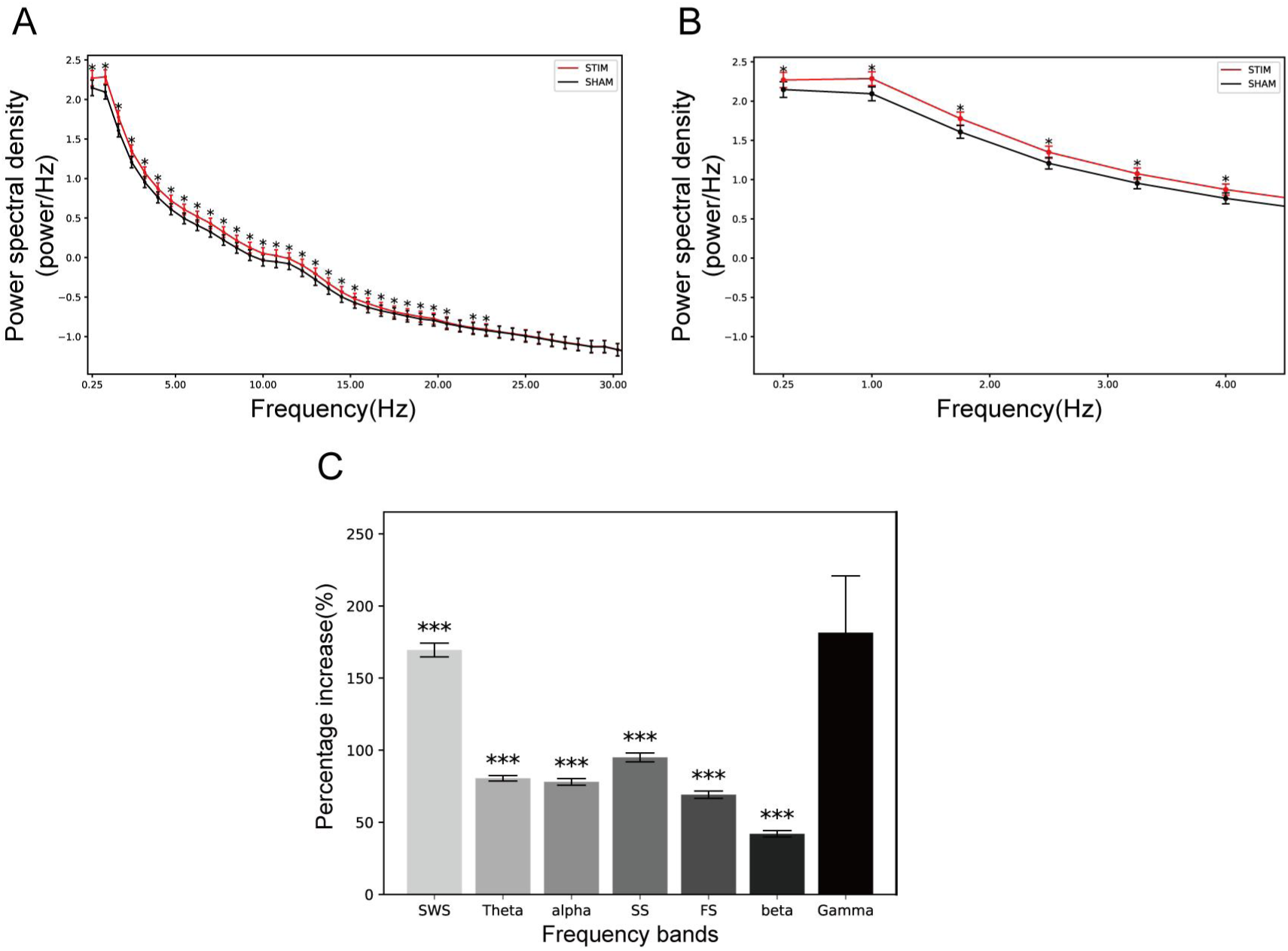
Spectral power of EEG activity during STIM and SHAM blocks. (A, B) Mean (±SEM) spectral power during 3.5s STIM and SHAM blocks for (A) frequencies up to 30 Hz and (B) frequencies up to 4 Hz, normalized to the MCP (0.25–30 Hz) across all blocks. Asterisk symbols indicate frequencies with significant differences (paired *t*-test, two-tailed). (C) Percentage increase in normalized power in the STIM compared to the SHAM conditions across seven frequency bands: SWA (0.5–4 Hz), theta (4–8 Hz), alpha (8–12 Hz), slow spindle (12–14 Hz), fast spindle (14–16 Hz), beta (16–20 Hz) and Gamma (30-100 Hz). Paired *t*-tests revealed significant differences across the first six bands. ***p<0.001, *p<0.05

### Event-related potentials (ERPs), Event-related Spectral Perturbations (ERSP) and Inter-trial Coherence (ITC) analysis

ERPs evoked by the first tone of each 2-tone block were averaged for STIM and SHAM conditions respectively (Figure 4A). In STIM condition, amplitude of slow oscillatory after tone delivery significantly enhanced relative to that in SHAM condition. Tones elicited a greater increase in the amplitude of slow oscillatory activity, with this effect tapering off after the second oscillation.

**Figure 4.**
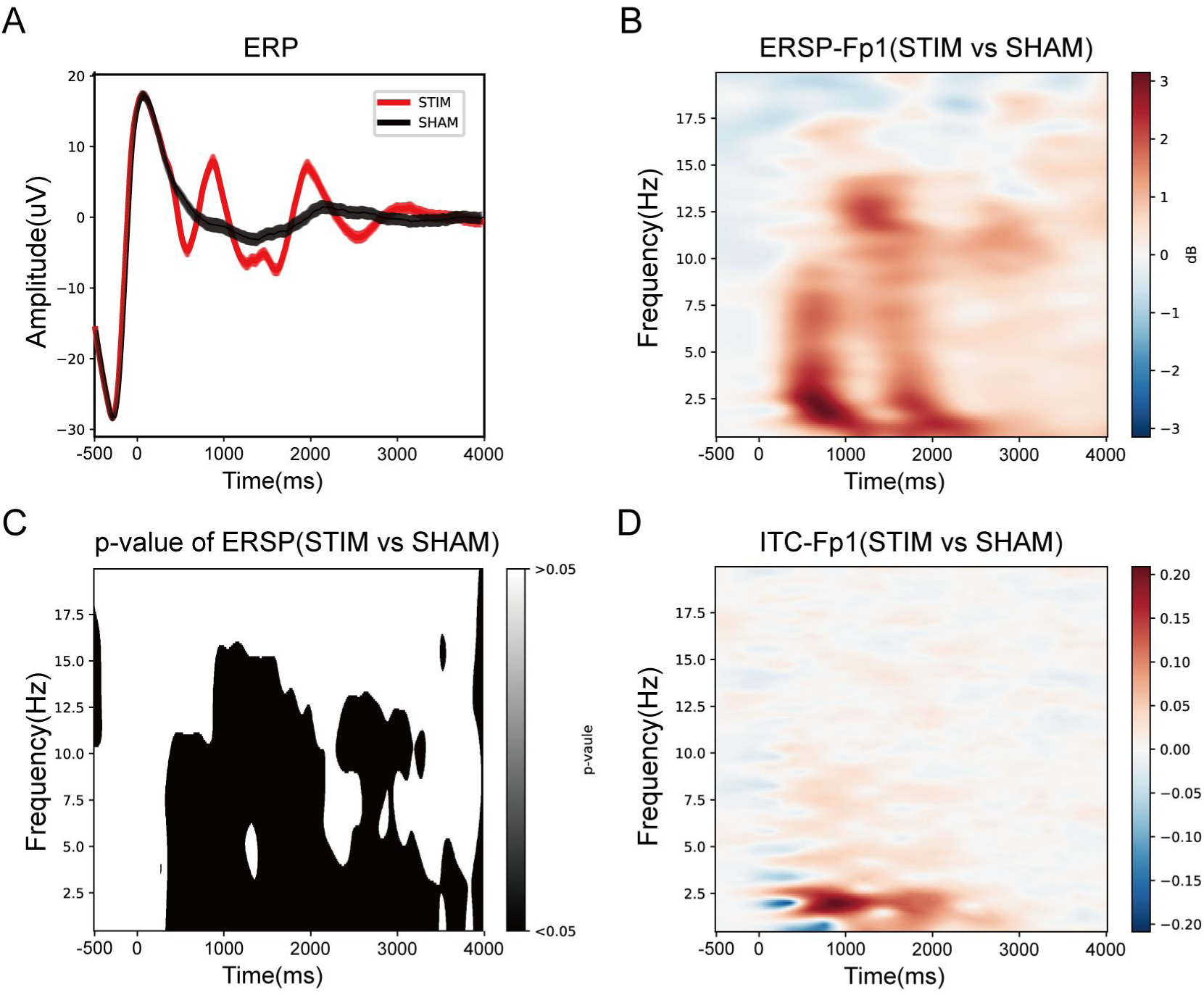
Averaged slow waves and Event-related spectral perturbation (ERSP). (A) Grand average event-related potentials (ERPs) for both the SHAM and STIM conditions(B) Event-related spectral perturbation (ERSP) and (C) inter-trial coherence (ITC) plots for STIM-SHAM at Fp1 locked to the onset of each block (time = 0). Increases in SWA (strong phase-locking across trials) and theta power were evoked, whilst brief increases in slow spindle activity were induced (weak phase-locking across trials). Panels (D) show p-values for (A).

To reveal the relationships of power changes among slow oscillations and other frequency bands, ERSP was plotted by calculating the changes of power spectra between STIM and SHAM conditions (figure 4B). This approach enabled us to identify induced (non-phase locked) activity that would have been diminished or canceled by signal averaging. We found that the power enhancement of theta (4∼8 Hz) and spindle activity (12∼16 Hz) were temporally confined to subsequent SWA up-state peaks, i.e., at approximately 1000 and 2000ms (arrows in figure 4B, C).

ITC plots further revealed that the phases of slow oscillations relative to tone stimulation in STIM conditions were highly consistent compared to that in SHAM condition (arrows in Figure 4D). This result indicated the phase lock between slow oscillations and tone stimulations.

As a conclusion, phase-locked tone stimulation significantly enhanced the slow oscillations, and the enhanced theta and spindle activity occurred at the SWA up-state peaks. Additionally, the second and third evoked slow oscillation also locked with tone stimulation.

## Discussion

In this study, we found that phase-locked tones with SWA peak significantly enhanced low-frequency brain oscillations, including SWA, theta, alpha, fast spindle, and slow spindle. Notably, the enhancement effect can be repeated for all participants by LANMAO system in home settings.

We used an “odd/even control” design to detect participants’ EEG changes in the home setting with unknow confounding factors. In contrast to prior research [18, 19], STIM and SHAM conditions in this study followed a within-subject design, employing an even-odd sequence. This design can minimize the sleep variations across participants and nights, thereby isolating the specific EEG effect of PLAS. It is important to highlight that the LANMAO device utilized in this study is designed for convenient home use, with the forehead patch weighing only 6 grams. This significantly reduces any potential interference caused by the device during sleep. Moreover, thanks to the precise N3 detection algorithm, only 1 out of the 6 subjects reported being awakened by auditory pulses before falling asleep or during the light sleep stage. These factors are of great significance for sleep-enhancing devices, as any negative impact on sleep quality may lead to reduced acceptance among patients, ultimately limiting the number of individuals who can benefit from such devices.

The device wireless transmission delay did not significantly affect the enhancement of SWA and spindle power.The precise timing of stimulation is crucial in acoustic stimulation. Out-of-phase stimulation, where pulses are emitted at the falling edge instead of the rising edge of the slow wave activity had no enhancing effects. A recent study demonstrated that stimulation applied at a random phase of the SWA resulted in an increase in SWA but a decrease in slow and fast spindle power, failing to improve overnight retention of word pairs.[20]. Our results indicate that phase-locked tone pulses, even when employed in a wireless transmission system with a ∼100 ms transmission delay, effectively target the upstate of slow oscillation. This finding is consistent with previous studies[21, 22]. These stimuli are precisely synchronized with the up-phase of SWA, leading to an increase in SWA and spindles, which are closely associated with memory consolidation.

According to the enhancement of alpha and beta power, the audio pulse may caused micro-arousals. While our primary focus in this study was to enhance SWA and spindles through acoustic stimulation, we unexpectedly observed concurrent increases in theta, alpha, and beta activity. These findings are slightly different with previous studies that primarily reported significant increases in SWA, and spindle power[3, 12, 23]. The implementation of a within-subject design, with closely temporally aligned STIM blocks and SHAM blocks, offers one plausible explanation for the observed disparities. This design choice facilitated the detection of subtle differences that could have been obscured by confounding variables across individuals and experimental sessions. Another potential explanation for the observed increase in theta power in our study relates to the association between delta and theta power and sleep homeostasis. These frequency bands have been found to exhibit heightened activity following periods of wakefulness and tend to decrease throughout the course of a sleep episode. Previous research has consistently demonstrated an elevation in both delta and theta power during recovery sleep following sleep deprivation, as well as across successive days of sleep restriction.[24]. Therefore, our study suggest that the acoustic stimulation may enhanced sleep intensity. Regarding the increase in slow and fast spindle power, our findings are consistent with prior research[7, 8, 15], Spindle activity plays a crucial role in memory consolidation within the thalamus[22, 25]. Therefore, the observed increase in spindle power supports the notion that memory consolidation of participants is enhanced. Regarding the enhancement observed in the alpha and beta bands, we hypothesize that it may be attributed to the micro-arousals induced by PLAS, based on the following cues: (1) Alpha bands have been observed to increase following auditory stimuli, such as K-complex-induced alpha bursts[26], and (2) beta activity typically increases during quiet wakefulness[27]. These findings suggest that the volume of 55 dB may disrupt the sleep process and require optimization, despite its widespread use in previous studies.[3, 12, 23, 28].

Several limitations should be noted. Firstly, the generalizability of our findings is limited to healthy, middle-aged subjects, warranting future research with diverse age groups and clinical populations, using larger sample sizes to enhance external validity[20, 29]. Furthermore, we did not conduct cognition tests in our study due to the limitations of self-administered tests in a home setting, which may introduce potential unreliability in the results. This limitation could be addressed in future research by employing more reliable assessment methods.Lastly, as a pilot study, we did not collect baseline sleep data for sleep structure analysis. Future studies will include baseline data to explore the effects of acoustic stimulation on sleep structures.

Enhancing slow-wave sleep in a home environment holds great significance because slow-wave sleep involved in various functions such as memory, immune response, and energy metabolism[1]. Slow-wave sleep facilitates memory consolidation [30, 31] and the clearance of beta-amyloid protein associated with Alzheimer’s disease, [32]. Restoring or enhancing slow-wave sleep at home may improve or delay cognitive impairments[28][33] and potentially mitigate immunological dysregulations [3]. Our study contributes to the translation of auditory slow-wave enhancement techniques from the lab to the home setting.

In conclusion, our study demonstrates that PLAS can also be effective in home settings, without the need for professional assistance. These findings open up new possibilities for the use of PLAS as a low-cost home-based intervention for sleep and cognitive function disorders.

## Data Availability

Data supporting the findings and computer code of this study are available from the corresponding author on reasonable request.

## Acknowledgment

We would like to thank the Shanghai QuanLan Technology team for their commitment to working on the LANMAO device.

